# Receipt of respiratory vaccines among patients with heart failure in a multicenter health system registry

**DOI:** 10.1101/2023.09.05.23295101

**Authors:** Anna Dermenchyan, Kristen R. Choi, Pooya R. Bokhoor, David J. Cho, Nina Lou A. Delavin, Chidinma Chima-Melton, Maria A. Han, Gregg C. Fonarow

**Affiliations:** Department of Medicine, Quality, University of California, Los Angeles, California; School of Nursing, University of California, Los Angeles, California; Department of Health Policy and Management, Fielding School of Public Health, University of California, Los Angeles, California; Department of Medicine, Division of Cardiology, University of California, Los Angeles, California; UCLA Faculty Practice Group, Office of Population Health and Accountable Care, University of California, Los Angeles, California

**Keywords:** heart failure classification, vaccine, health system registry

## Abstract

**Background:** Heart failure affects people of all ages and is a leading cause of death for both men and women in most racial and ethnic groups in the United States. Infections are common causes of hospitalizations in heart failure, with respiratory infections as the most frequent diagnosis.

Vaccinations provide significant protection against preventable respiratory infections. Despite being an easily accessible intervention, prior studies suggest vaccines are underused in patients with heart failure.

**Methods:** An observational study of 7341 adults with heart failure was conducted using data from an integrated, multicenter, academic health system in Southern California from 2019 to 2022. Logistic regression models were used to determine the rates of influenza, pneumococcal, and COVID-19 vaccination among a population of patients with heart failure (heart failure preserved ejection fraction [HFpEF], heart failure mildly reduced ejection fraction [HFmrEF], heart failure reduced ejection fraction [HFrEF], and heart failure unspecified ejection fraction [HFuEF]) and identify whether heart failure phenotype is associated with vaccination status.

**Results:** Vaccination rates varied between influenza, pneumococcal, and COVID-19 vaccines. Of the three respiratory vaccines, 54.5% of patients had received an influenza vaccine, 74.7% had received a pneumococcal vaccine, and 81.3% had received a COVID-19 vaccine. There were no sex-based differences by vaccination status. Patients with HFpEF and HFmrEF had the highest vaccination levels in all three vaccine groups. In adjusted models, patients with HFpEF had higher odds of being vaccinated for influenza (aOR=1.34, 95% CI=1.19-1.53), pneumococcal (aOR=1.28, 95% CI=1.10-1.48), and COVID-19 (aOR=1.25, 95% CI=1.07-1.47) compared to HFuEF patients. Patients with HFrEF had lower odds of being vaccinated for pneumococcal (aOR=0.81, 95% CI=0.70-0.93) than patients with HFuEF.

**Conclusions:** Patients with HFrEF had the lowest levels of respiratory vaccination compared to other specified heart failure categories. Interventions are needed to increase vaccination education and offerings, especially to patients with HFrEF.

## Introduction

Heart failure is associated with considerable morbidity, mortality, and cost. Infections are common causes of hospitalizations in heart failure, with respiratory infections as the most frequent diagnosis [1, 2]. Respiratory infections can cause significant disease and poor outcomes in patients with heart failure. Influenza, pneumococcal, and COVID-19 vaccines can help protect against significant morbidity or mortality in vulnerable heart failure patients and reduce the incidence of common respiratory infections [3-6]. Multiple studies have shown the benefits of vaccination for preventing and managing heart disease [3, 7-12]. Despite being an easily accessible intervention, vaccinations are underutilized and understudied in patients with heart failure.

Patients with heart failure are at a higher risk of severe complications from respiratory illnesses [2, 13]. Influenza, pneumonia, and COVID-19 infections can trigger arrhythmias, acute coronary syndromes, and acute exacerbations of underlying heart failure [14]. Vaccinations provide the best protection against preventable respiratory disease [3]. A large, recent trial conducted in low- and middle-income countries confirmed that influenza vaccination could reduce the risk of hospitalization and pneumonia in patients with symptomatic heart failure [15, 16]. However, prior studies have found low levels of vaccination in the heart failure population. In a study of 313761 patients hospitalized at centers participating in the Get With The Guidelines-Heart Failure (GWTG-HF) registry from 2012 to 2017 [8], nearly 1 in 3 patients hospitalized with heart failure were not vaccinated for influenza or pneumococcal, and vaccination rates did not improve across the five-year study period. The hospitals with higher vaccination rates performed well in other heart failure quality of care measures [7, 8].

Guideline‐recommended therapies are vital in improving outcomes and preventing recurrent cardiovascular events [17, 18]. Due to a lack of randomized clinical trials, heart failure guidelines from the American Heart Association and American College of Cardiology Foundation (AHA/ACCF), Heart Failure Society of America (HFSA), and European Society of Cardiology (ESC) do not cover vaccination recommendations in detail. The U.S. Preventive Services Task Force (USPSTF) and Advisory Committee on Immunization Practices (ACIP) remain vague on specific vaccine recommendations but make general recommendations for chronic heart, lung, liver, diabetes, and smoking [5]. Nonetheless, there is consensus in the scientific community to provide influenza, pneumococcal, and COVID-19 vaccines to all patients with heart failure in the absence of contraindications [19, 20]. Given limited evidence on respiratory vaccines among patients with heart failure, especially for the newer COVID-19 vaccine, the aim of this study was to investigate levels of influenza, pneumococcal, and COVID-19 vaccination among a heart failure patient population and whether heart failure phenotype was associated with vaccination status.

## Methods

### Design and Data

This observational study used data from an integrated, multicenter academic health system from 2019 to 2022. The database included all known adult patients with heart failure diagnoses on their Electronic Health Record (EHR) problem list. The problem list is updated by clinicians who are members of the patient’s care team in ambulatory and inpatient settings. The registry includes data about patient demographics, clinical and social characteristics, treatment background, and clinician type. The UCLA Institutional Review Board provided a determination of exempt status for this study as an analysis of existing de-identified data.

### Sample and Setting

The study cohort included 7341 adults over 18 years of age with heart failure. The health system in Southern California is comprised of four hospitals and over 250 ambulatory care clinics with more than 2.5 million patient visits and 100000 hospital admissions annually. All individuals listed in the dataset were eligible for inclusion in the study sample, and the entire cohort was used for the analysis.

### Outcome Variable: Vaccination Status

Vaccination status data were populated into the patient registry from the patient’s immunization record in the EHR. Vaccination status data included within-health system and external data sources. External immunization records were populated from the California Immunization Registry (CAIR2) and Epic Care Everywhere (Epic Systems Corporation, Inc. Verona, WI). The CAIR2 is a secure, confidential, statewide computerized immunization information system for California residents. Epic Care Everywhere allows health systems to share medical records with other health systems using the same EHR. Patients were considered vaccinated for each vaccine type when the following conditions were met:

1. Influenza vaccine: documentation of immunization type (42 total) or professional billing charge (20 total) in the EHR during the influenza vaccination period from August of the previous year to March of the current year.
2. Pneumococcal vaccine: documentation of one dose of pneumococcal polysaccharide vaccine 23-valent (Pneumovax) or pneumococcal conjugate vaccine 13-valent (Prevnar 13) (no time specified). The pneumococcal conjugate vaccine 20-valent (Prevnar 20) was not captured in the EHR during the study period; it was added two months later.
3. COVID-19 vaccine: documentation of two doses for the mRNA manufacturers (Pfizer-BioNTech, Moderna), one dose for Johnson and Johnson, or one dose for unspecified (no time specified).

### Exposure Variable: Heart Failure Category

Heart failure type was classified into four categories: heart failure with preserved ejection fraction (HFpEF, ejection fraction [EF] ≥50%), heart failure with mildly reduced ejection fraction (HFmrEF, EF 41-49%), heart failure with reduced ejection fraction (HFrEF, EF ≤40%), and heart failure with unspecified ejection fraction (HFuEF, EF unspecified) [21, 22]. Over 2000 ICD-10-CM codes/subsets are associated with a heart failure diagnosis, comprising approximately 3% of total ICD-10 codes. In the health system heart failure registry, these various ICD-10-CM diagnosis codes were populated from the EHR’s problem list into the registry and categorized into the four heart failure groups. To improve the accuracy of the problem list, clinicians were notified through a Best Practice Alert (BPA) in the EHR when a heart failure diagnosis may be erroneously missing from the problem list based on documentation in the medical history, one or more billing codes, EF value, or prescriptions of heart failure medications. Within the healthcare system, there was an active push to refine generic heart failure diagnoses to more specific ones as problems became more defined during the course of patient treatment. If multiple heart failure diagnoses were on the problem list, the algorithm selected the diagnosis based on the lowest EF on file in the last three years. Based on this logic, there was no overlap between the heart failure categories.

### Covariates

Covariates were demographic, social, and health characteristics that might influence the relationship between exposures and outcomes. These included patient gender (male/female), age (<65 years, ≥65 years), race/ethnicity (Asian, Black, Hispanic, White, Other), primary language (English, non-English), Accountable Care Organization status (ACO), insurance type (Commercial, Medicaid, Medicare, Managed Care, Other), and a Social Vulnerability Index (SVI) score. SVI measured four domains of social vulnerability from United States Census data, including (1) socioeconomic status, (2) household composition and disability, (3) minority status and language, and (4) housing and transportation [23]. Higher scores denote higher social vulnerability. We measured patient comorbidity characteristics, including the presence of chronic kidney disease (CKD), diabetes mellitus, hypertension, and ischemic heart disease. Finally, we assessed whether the patient was registered with a primary care physician, cardiologist, or both in the health system.

### Statistical Analysis

RStudio was used for statistical analysis. Descriptive statistics and frequencies were used to characterize the sample on all analytic variables. Chi square tests were used to compare sample differences by patient and clinician characteristics and vaccination status. We estimated odds of vaccination by heart failure type in three logistic regression models, first assessing the bivariate relationship between the exposure and outcome and then adjusting for all covariates listed above. For both unadjusted and adjusted models, we estimated separate models for each vaccination outcome.

## Results

Patient and clinician characteristics of the heart failure population by vaccination status are presented in Table 1. The sample was primarily male (55.2%, n=4049), aged 65 years or older (74.5%, n=5467), self-identified as White (52.9%, n=3886), and insured by Medicare (66.1%, n=4851). Thirty-one percent (n=2256) had an SVI score of medium to high. Sixty percent of patients had a registered primary care physician (n=4430), while 52.6% had a registered cardiologist (n=3863) and 36.4% had both clinician types (n=2675). Of the total sample, 37.7 % (n=2764) were patients with HFpEF, 2.7% (n=198) were heart failure with HFmrEF, 28.9% (n=2127) were patients with HFrEF, and 30.7% (n=2252) were patients with HFuEF (Figure 1). Patient comorbidity characteristics included 30% CKD (n=2205), 32.1% diabetes mellitus (n=2356), 74.8% hypertension (n=5494), and 21% ischemic heart disease (n=1545).

**Table 1.**
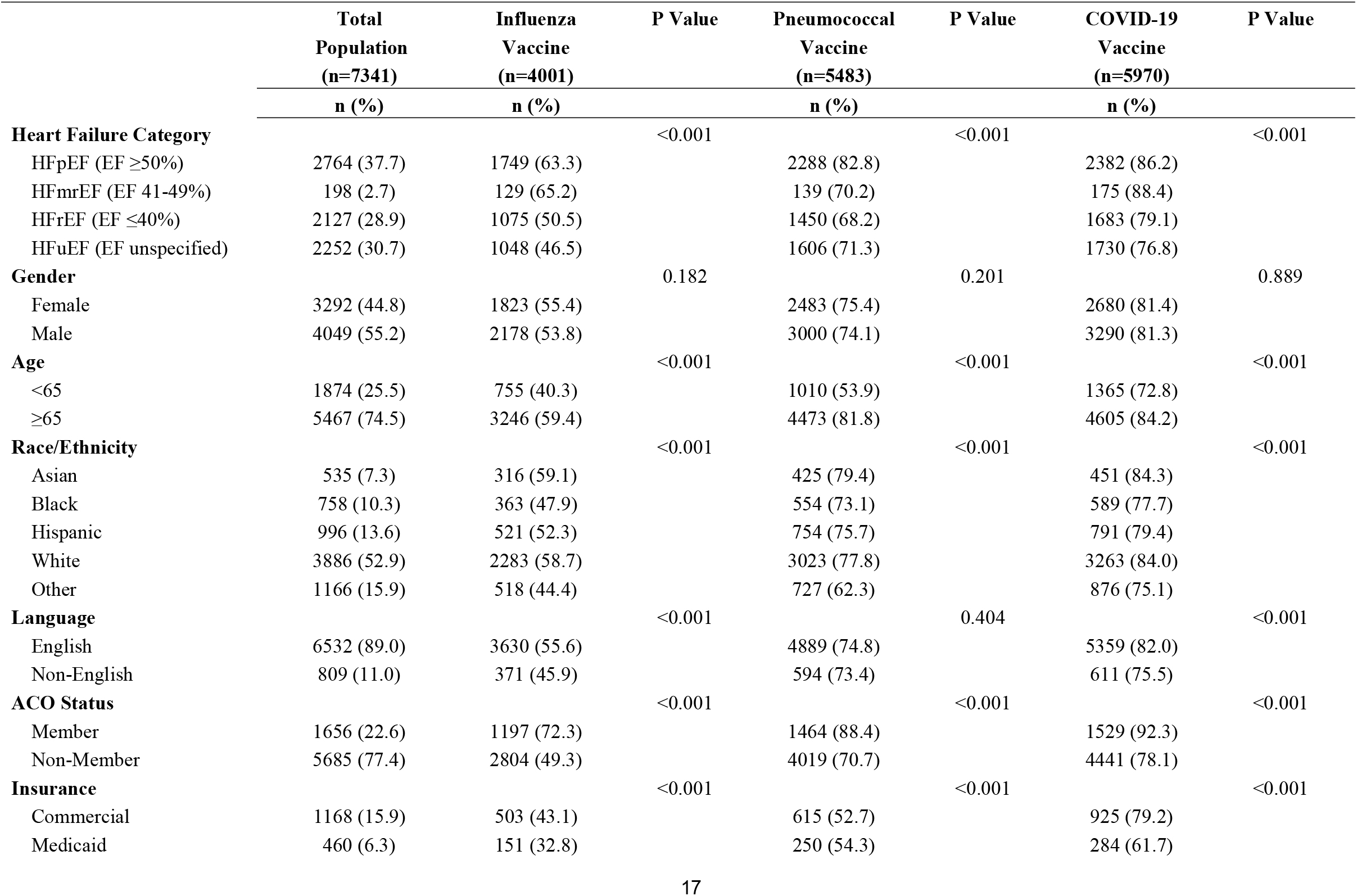
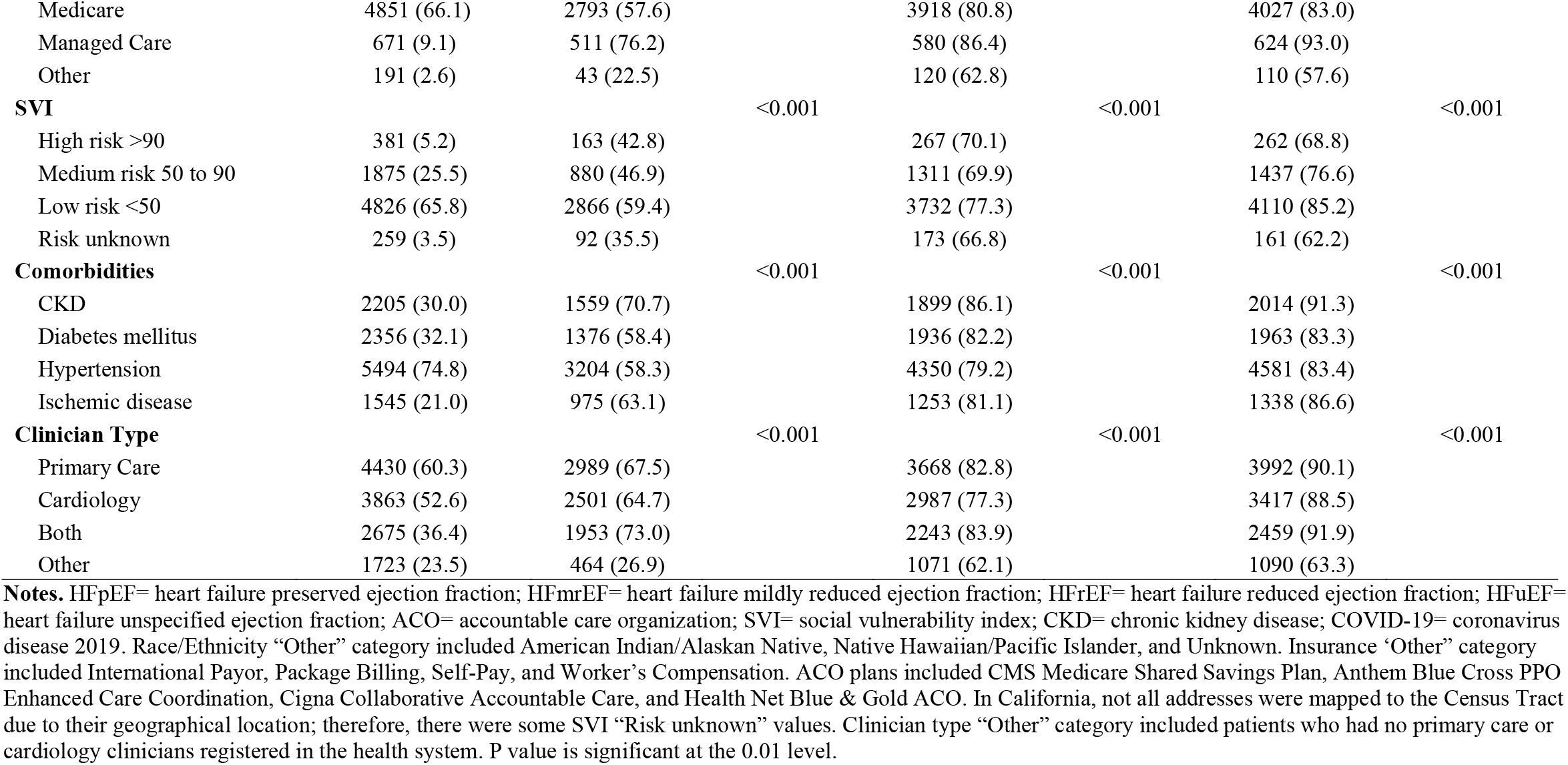
Patient and clinician characteristics by influenza, pneumococcal, and COVID-19 vaccination status.

**Figure 1:**
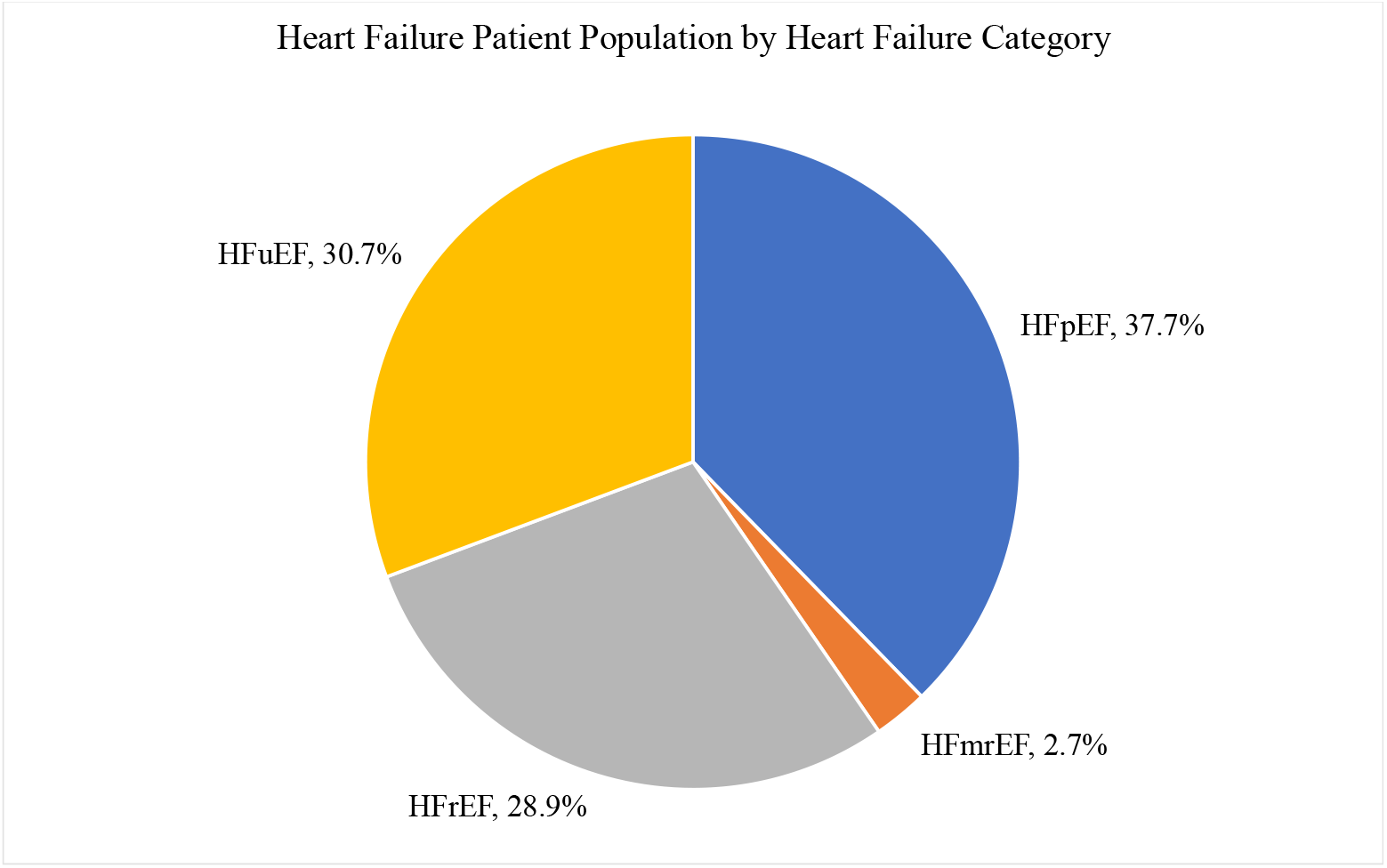
Heart failure patient population by heart failure category. **Legend**: Proportion of adult patients with each of four heart failure types in a sample of 7341 patients with heart failure derived from a health system Heart Failure Registry in Southern California from 2019 to 2022. HFpEF= heart failure preserved ejection fraction; HFmrEF= heart failure mildly reduced ejection fraction; HFrEF= heart failure reduced ejection fraction; HFuEF= heart failure unspecified ejection fraction.

There were no sex-based differences seen in vaccination status. An estimated 54.5% of patients (n=4001) received an influenza vaccine, 74.7% (n=5483) received a pneumococcal vaccine, and 81.3% (N=5970) received a COVID vaccine (Figure 2). In bivariate tests, there were significant differences in vaccination status by heart failure category for all three vaccines (p<0.001). In multiple logistic regression models, patients with HFpEF (aOR=1.34, 95% CI=1.19-1.53) and HFmrEF (aOR=1.40, 95% CI=1.01-1.95) had higher odds of being vaccinated for influenza compared to patients with HFuEF. Likewise, patients with HFpEF had higher odds of being vaccinated for pneumococcal (aOR=1.28, 95% CI=1.10-1.48), and patients with HFrEF had lower odds of being vaccinated for pneumococcal compared to patients with HFuEF (aOR=0.80, 95% CI=0.70-0.93). Patients with HFpEF had higher odds of being vaccinated for COVID-19 than those with HFuEF (aOR=1.25, 95% CI=1.07-1.47) (Table 2).

**Table 2.** Odds of vaccination by heart failure category.

| Predictors | Influenza Vaccine |  | Pneumococcal Vaccine |  | COVID-19 Vaccine |  |
| --- | --- | --- | --- | --- | --- | --- |
|  | OR (95% CI) | aOR (95% CI) | OR (95% CI) | aOR (95% CI) | OR (95% CI) | aOR (95% CI) |
| **Heart Failure Category** (reference: HFuEF [EF unspecified]) |  |  |  |  |  |  |
| HFpEF (EF ≥50) | 1.98 (1.77-2.22) | 1.34 (1.19-1.53) | 1.93 (1.69-2.21) | 1.28 (1.10-1.48) | 1.88 (1.63-2.18) | 1.25 (1.07-1.47) |
| HFmrEF (EF 41-49%) | 2.15 (1.59-2.92) | 1.40 (1.01-1.95) | 0.95 (0.69-1.31) | 0.76 (0.54-1.09) | 2.30 (1.50-3.68) | 1.34 (0.86-2.20) |
| HFrEF (EF ≤40%) | 1.17 (1.04-1.32) | 1.00 (0.87-1.14) | 0.86 (0.76-0.98) | 0.81 (0.70-0.93) | 1.14 (0.99-1.32) | 0.94 (0.81-1.10) |

**Notes.** Three unadjusted and adjusted multiple logistic regression models estimating odds of vaccination for influenza, pneumococcal, and COVID-19 vaccines by heart failure category. Adjusted models included covariates for gender, age, race/ethnicity, insurance type, whether the patient was a member of an Accountable Care Organization (ACO), primary language, Social Vulnerability Index (SVI) score, clinician type, and number of comorbidities. In this analysis, N=7341 represents patients with heart failure from 2019 to 2022 in the health system Heart Failure Registry. HFpEF= heart failure preserved ejection fraction; HFmrEF= heart failure mildly reduced ejection fraction; HFrEF= heart failure reduced ejection fraction; HFuEF= heart failure unspecified ejection fraction; EF= Ejection Fraction; OR= odds ratio; CI= confidence interval.

**Figure 2:**
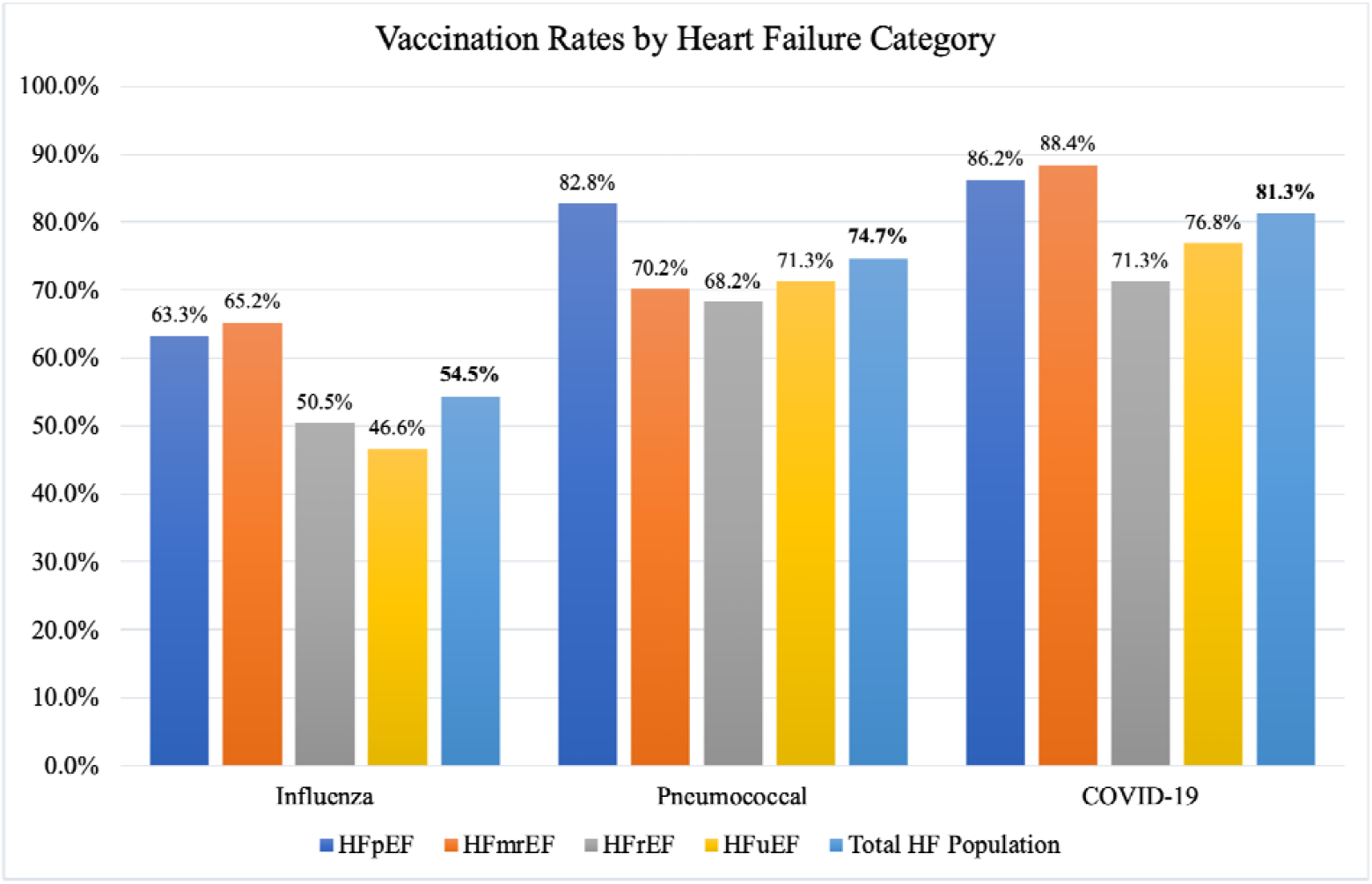
Vaccination rates for influenza, pneumococcal, and COVID-19 vaccines by heart failure category. **Legend:** Percentage of adult patients who received respiratory vaccines (influenza, pneumococcal, COVID-19) by heart failure type. The sample includes 7341 adult patients with heart failure derived from a health system Heart Failure Registry in Southern California from 2019 to 2022. HFpEF= heart failure preserved ejection fraction; HFmrEF= heart failure mildly reduced ejection fraction; HFrEF= heart failure reduced ejection fraction; HFuEF= heart failure unspecified ejection fraction.

## Discussion

There have been very few studies examining the receipt of common respiratory vaccines concurrently among patients with heart failure, especially investigating the receipt of COVID-19 vaccines. In this study of adults with heart failure, we found that individuals with HFrEF had the lowest rates of respiratory vaccination that could prevent infections and hospitalizations. Patients with HFpEF had the highest odds of receiving all three vaccine types, adjusting for demographic and social factors and patient comorbidities. Vaccination rates for the three vaccines were comparable to the general public [20], but there were significant gaps in vaccination for patients with HFrEF who could benefit most from this preventive intervention.

Of the three respiratory vaccines, the vaccination rates were the lowest for the influenza vaccine (54.5%). The influenza vaccine has been available since 1945, but the public interest in receiving this vaccine is low. A 2021 survey found that in the general U.S. public, only 56% (N=631) of adults wanted to receive an influenza vaccine, and among those who were unsure, 39% did not believe flu vaccines worked very well [24]. However, survey participants also believed that healthcare providers were the primary and most trusted source of information about influenza and influenza vaccination, suggesting that healthcare providers play an important role in vaccine uptake. The Centers for Disease Control and Prevention recommends assessing the vaccination status of patients and addressing misconceptions about vaccines at all clinical encounters [19]. Incorporating tailored patient education on preventive practices like annual flu shots as part of patient self-management, especially for patients with more severe heart failure, may increase vaccine uptake.

Pneumococcal pneumonia infections are one of the leading causes of heart failure admission [25, 26]. In this study, patients with HFrEF had the lowest levels of pneumococcal vaccination and lower odds of vaccination than other heart failure types. Some possible reasons might be that these patients were managed mainly by cardiologists focused on advanced therapies than closing preventive care gaps. In addition, some cardiology clinics did not have the workflows to provide respiratory vaccinations to patients during their clinic visits. Prior studies have found a high incidence of pneumonia among patients with heart failure, even those with HFpEF [27]. Meta-analyses have also found that in the general adult population, pneumococcal vaccination is associated with a decreased risk of cardiovascular events and myocardial infarction in all age groups [4]. Since much of the clinical management of patients with HFpEF is primarily directed toward treating associated conditions [17], vaccination in this population may produce more favorable outcomes.

Underlying heart disease is associated with an increased risk for in-hospital death among patients hospitalized with COVID-19 [28]. Of the three vaccines in this study, the vaccination rates were the highest for the COVID-19 vaccine (81.3%). At the beginning of the COVID-19 pandemic, there was a great deal of media attention to COVID-19 hospitalization and mortality rates for those with high-risk medical conditions, and initially, COVID-19 vaccines became available earlier for individuals with high-risk medical conditions, including heart failure [24]. Efforts to increase COVID-19 vaccination in this population may explain why COVID-19 vaccination levels were high in our sample. However, there were still significant vaccination differences among the four heart failure categories, with those with HFrEF or HFuEF having the lowest levels of COVID-19 vaccination. As such, there is a need for targeted communication and outreach to patients with HFrEF.

This study suggests an urgent need to reach patients with heart failure with respiratory vaccines, especially the influenza vaccine, which had the lowest rates of uptake in our study. Although education on the need for respiratory vaccines in clinical encounters can improve care for some patients, a systems approach could be more effective at improving the health of the entire population and reducing vaccine inequities. Clinicians should utilize both primary care and cardiology encounters to assess the vaccination status of patients and address patient hesitancy and vaccine side effects and long-term effects. We also recommend integration with external state vaccine registries to ensure up-to-date and accurate capture of vaccination status, provide vaccines within the visit, and offer referrals to retail pharmacy if needed. Clinicians may also benefit from tracking population-level data at the provider and clinic levels to set organizational benchmarks for vaccination, make data transparent, and provide incentives to increase vaccination rates. Future research could evaluate patient and clinician factors that influence vaccine rates. The 2022 ACC/AHA/HFSA heart failure guidelines provide a Class 2a, B-NR recommendation that vaccinating against respiratory illnesses is reasonable to reduce mortality [29]. In the future, it may be beneficial for such clinical guidelines to make specific recommendations for respiratory vaccines.

## Limitations

There are strengths and limitations to this study to consider in interpreting the findings. Our study used a large, population-based cohort of adults with heart failure derived from a multicenter integrated academic health system. Validity testing of the data was conducted by a team of primary care and cardiology clinicians, quality specialists, and computer programmers and analysts during a six-month period prior to data analysis. However, a clinician had to actively include heart failure on the EHR’s problem list for the patient to be included, and thus it is possible that this dataset does not capture all patients in the health system with heart failure. We had complete vaccine outcomes data pulled from multiple sources, including external sources of CAIR2 and Epic Care Everywhere. Our study did not capture patients who received Prevnar 20 for the pneumococcal vaccine, which was added to the health system EHR after the time of the study. In addition, the COVID-19 boosters were not counted for numerator compliance. At the time of the study, hospitalization and mortality data were unavailable for the patient population. Therefore, additional analysis could not be done to link vaccination status to outcomes data. In addition, vaccination status in relation to guideline-directed medical therapy compliance was not measured. These areas are of interest for future studies. Despite these limitations, our findings can inform ways to implement population-level interventions to improve vaccination outcomes among patients with heart failure.

## Conclusion

Heart failure is a complex chronic disease to manage, but preventive care practices like vaccinations can improve patient outcomes and reduce the risk of infections, hospitalization, and mortality. This study found differences in respiratory vaccination rates by heart failure categories and highlights care gaps that can be addressed through population-level strategies to increase vaccination. Because patients with HFrEF who could potentially benefit most from respiratory vaccines are least likely to receive them, it is important for individual clinicians as well as health systems to develop strategies to improve use and overcome heart failure vaccine differences.

## Data Availability

This study used internal data from UCLA Health's Heart Failure Registry.

## Acknowledgments

We want to acknowledge the UCLA Department of Medicine Quality and Cardiology Division’s dedication and hard work in supporting quality improvement efforts for the heart failure patient population.

## References

1. Fonarow, G.C., et al., Factors identified as precipitating hospital admissions for heart failure and clinical outcomes: findings from OPTIMIZE-HF. Arch Intern Med, 2008. 168(8): p. 847–54.

2. Alon, D., et al., Predictors and outcomes of infection-related hospital admissions of heart failure patients. PLoS One, 2013. 8(8): p. e72476.

3. Girerd, N., et al., Vaccination for Respiratory Infections in Patients with Heart Failure. J Clin Med, 2021. 10(19).

4. Marra, F., et al., The protective effect of pneumococcal vaccination on cardiovascular disease in adults: A systematic review and meta-analysis. Int J Infect Dis, 2020. 99: p. 204–213.

5. Bhugra, P., et al., Determinants of Influenza Vaccine Uptake in Patients With Cardiovascular Disease and Strategies for Improvement. J Am Heart Assoc, 2021. 10(15): p. e019671.

6. Gotsman, I., et al., Influenza Vaccination and Outcome in Heart Failure. Am J Cardiol, 2020. 128: p. 134–139.

7. Bhatt, A.S., et al., Can Vaccinations Improve Heart Failure Outcomes?: Contemporary Data and Future Directions. JACC Heart Fail, 2017. 5(3): p. 194–203.

8. Bhatt, A.S., et al., Vaccination Trends in Patients With Heart Failure: Insights From Get With The Guidelines-Heart Failure. JACC Heart Fail, 2018. 6(10): p. 844–855.

9. Fountoulaki, K., et al., Beneficial Effects of Vaccination on Cardiovascular Events: Myocardial Infarction, Stroke, Heart Failure. Cardiology, 2018. 141(2): p. 98–106.

10. Rodrigues, B.S., et al., Influenza vaccination in patients with heart failure: a systematic review and meta-analysis of observational studies. Heart, 2020. 106(5): p. 350–357.

11. Vardeny, O., et al., Influenza Vaccination in Patients With Chronic Heart Failure: The PARADIGM-HF Trial. JACC Heart Fail, 2016. 4(2): p. 152–158.

12. Ciszewski, A., Cardioprotective effect of influenza and pneumococcal vaccination in patients with cardiovascular diseases. Vaccine, 2018. 36(2): p. 202–206.

13. Drozd, M., et al., Infection-Related Hospitalization in Heart Failure With Reduced Ejection Fraction: A Prospective Observational Cohort Study. Circ Heart Fail, 2020. 13(5): p. e006746.

14. Rojulpote, C., et al., COVID-19 and the Heart. Colomb Med (Cali), 2020. 51(2): p. e4320.

15. Kumbhani, D.J. Influenza Vaccine to Prevent Adverse Vascular Events - IVVE. 2022 [cited 2022 September 10, 2022]; Available from: https://www.acc.org/latest-in-cardiology/clinical-trials/2022/04/02/15/50/ivve.

16. Loeb, M., et al., Randomized controlled trial of influenza vaccine in patients with heart failure to reduce adverse vascular events (IVVE): Rationale and design. Am Heart J, 2019. 212: p. 36–44.

17. Yancy, C.W., et al., 2013 ACCF/AHA guideline for the management of heart failure: executive summary: a report of the American College of Cardiology Foundation/American Heart Association Task Force on practice guidelines. Circulation, 2013. 128(16): p. 1810–52.

18. Yancy, C.W., et al., 2017 ACC/AHA/HFSA Focused Update of the 2013 ACCF/AHA Guideline for the Management of Heart Failure: A Report of the American College of Cardiology/American Heart Association Task Force on Clinical Practice Guidelines and the Heart Failure Society of America. Circulation, 2017. 136(6): p. e137–e161.

19. Centers for Disease Control and Prevention. Strategies for Increasing Adult Vaccination Rates. 2021; Available from: https://www.cdc.gov/vaccines/hcp/adults/for-practice/increasing-vacc-rates.html.

20. Centers for Disease Control and Prevention. Vaccination Coverage among Adults in the United States, National Health Interview Survey. 2022; Available from: https://www.cdc.gov/vaccines/imz-managers/coverage/adultvaxview/pubs-resources/vaccination-coverage-adults-2019-2020.html.

21. Ponikowski, P., et al., 2016 ESC Guidelines for the diagnosis and treatment of acute and chronic heart failure: The Task Force for the diagnosis and treatment of acute and chronic heart failure of the European Society of Cardiology (ESC)Developed with the special contribution of the Heart Failure Association (HFA) of the ESC. Eur Heart J, 2016. 37(27): p. 2129–2200.

22. Savarese, G., et al., Heart failure with mid-range or mildly reduced ejection fraction. Nat Rev Cardiol, 2022. 19(2): p. 100–116.

23. Lewis, B.E.F.E.W.G.E.J.H.J.L.H.a.B., A Social Vulnerability Index for Disaster Management. Journal of Homeland Security and Emergency Management, 2011. 8(1).

24. National Foundations for Infectious Diseases. 2021 National Survey: Attitudes about Influenza, Pneumococcal Disease, and COVID-19. 2021; Available from: https://www.nfid.org/2021-national-survey-attitudes-about-influenza-pneumococcal-disease-and-covid-19/.

25. Mor, A., et al., Chronic heart failure and risk of hospitalization with pneumonia: a population-based study. Eur J Intern Med, 2013. 24(4): p. 349–53.

26. Thomsen, R.W., et al., The impact of pre-existing heart failure on pneumonia prognosis: population-based cohort study. J Gen Intern Med, 2008. 23(9): p. 1407–13.

27. Shen, L., et al., Incidence and Outcomes of Pneumonia in Patients With Heart Failure. J Am Coll Cardiol, 2021. 77(16): p. 1961–1973.

28. Mehra, M.R., et al., Cardiovascular Disease, Drug Therapy, and Mortality in Covid-19. N Engl J Med, 2020. 382(25): p. e102.

29. Heidenreich, P.A., et al., 2022 AHA/ACC/HFSA Guideline for the Management of Heart Failure: A Report of the American College of Cardiology/American Heart Association Joint Committee on Clinical Practice Guidelines. Circulation, 2022. 145(18): p. e895–e1032.

